# Survival With a Cost: Increased Long-Term Coronary Artery Disease Risk After Adjuvant Fluoropyrimidine Chemotherapy in Colorectal Cancer Survivors

**DOI:** 10.1101/2025.08.29.25334733

**Authors:** Ivana Iveljić, Lejla Alidžanović Nurkanović, Nay Aung, Dunja Aksentijevic

## Abstract

**Background:** Fluoropyrimidine-based chemotherapy, including 5-fluorouracil (5-FU) and capecitabine, is a cornerstone of adjuvant treatment in non-metastatic colorectal cancer (CRC). While acute cardiotoxicity is recognized, the long-term impact on coronary artery health remains poorly understood. This study aimed to determine whether CRC survivors treated with fluoropyrimidines exhibit a higher burden of coronary artery disease (CAD) years after therapy completion.

**Methods:** In this cross-sectional, single-center study, CRC survivors five to seven years post-adjuvant 5-FU/capecitabine therapy (ChemT group) undergoing elective coronary angiography, were compared with age- and sex-matched cancer-free controls undergoing elective coronary angiography (n=45/group). Clinical, laboratory, echocardiographic, and invasive angiographic parameters were systematically evaluated. The primary endpoint was the presence and anatomical distribution of significant CAD.

**Results:** Despite fewer anginal symptoms, ChemT patients had a significantly higher prevalence of CAD in the proximal left anterior descending artery (24% vs 2%, *P*=0.004) and proximal right coronary artery (13% vs 0%, *P*=0.026). Overall, 44% of ChemT patients required percutaneous coronary intervention versus 16% of controls (*P*=0.006), with a greater number of stents implanted in the ChemT group. Notably, the ChemT cohort also demonstrated elevated LDL cholesterol, hepatic transaminases, and abnormal hematologic indices, suggestive of long-term metabolic and vascular stress. Echocardiography revealed no significant impairment in ejection fraction but altered left ventricular geometry in the ChemT group compared to controls.

**Conclusions:** CRC survivors treated with fluoropyrimidine-based chemotherapy exhibit a significantly elevated burden of anatomically high-risk coronary artery disease, particularly in proximal segments with poor prognostic implications. These findings highlight a silent but clinically significant cardiovascular risk in long-term cancer survivorship and support the need for proactive cardiac surveillance strategies in this population.

**Graphical Abstract:** 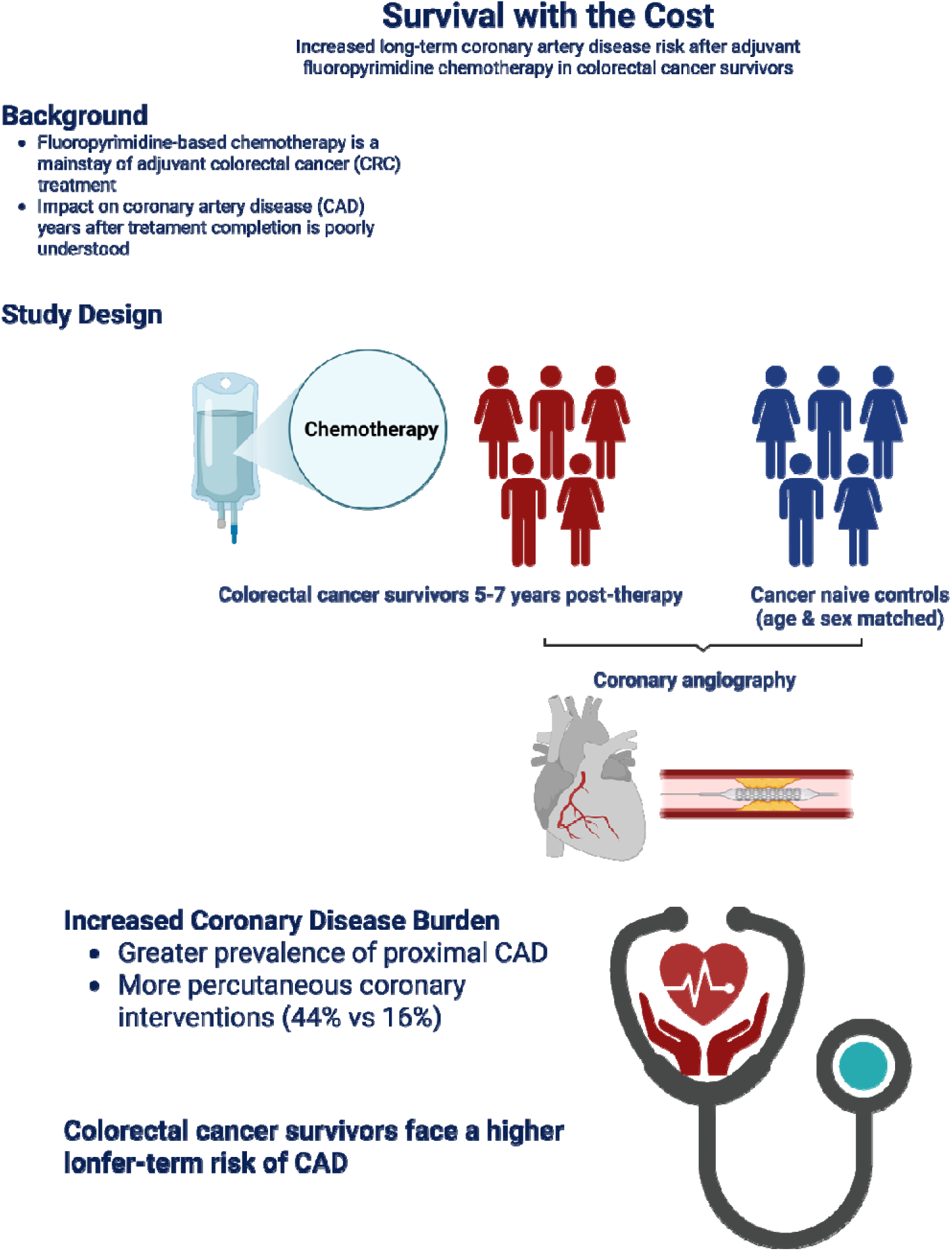

## Introduction

Colorectal carcinoma (CRC) accounts for approximately 10% of all malignancies worldwide and is the third most common cause of cancer-related death, responsible for over 600,000 deaths annually^1^. Despite progress in early detection and treatment, the global burden of CRC remains substantial. Five-year survival rates vary widely depending on disease stage and patient demographics, ranging from 30% to 60% in women and 28% to 57% in men^2^.

For patients with non-metastatic CRC, surgical resection remains the cornerstone of treatment, offering the potential for cure and providing essential histopathological staging^3^. In high-risk stage II and all stage III cases, adjuvant chemotherapy is the standard of care. Fluoropyrimidine-based regimens, often combined with oxaliplatin, are commonly used to reduce recurrence risk and improve survival outcomes^4^. Among fluoropyrimidines, intravenous 5-fluorouracil (5-FU) and its oral prodrug capecitabine converted to 5-FU in tumour tissues are the most widely utilized agents.

Despite therapeutic advances, CRC remains a leading cause of death within the first five years after diagnosis^5^. For those who survive beyond this critical period, non-cancer causes of death become increasingly relevant. Notably, cardiovascular disease (CVD) has emerged as the predominant cause of long-term mortality in CRC survivors^6^. Strikingly, the incidence of cardiovascular disease reaches ∼57% among older CRC survivors, more than double the ∼22% observed in the general population of similar age underscoring the significant cardiovascular burden in this population^7^.

Cardiotoxicity is a recognized complication of fluoropyrimidine therapy, affecting an estimated 1% to 4% of patients^8^. These cardiotoxic manifestations are observed during the administration of therapy and represent transient, short-term effects ranging from chest pain and electrocardiographic changes to coronary vasospasm, and even acute myocardial infarction^8^. Preclinical models have identified ischemic injury and direct cellular toxicity as potential mechanisms driving these acute events^9^. However, the long-term cardiovascular effects of fluoropyrimidine-based chemotherapy remain incompletely understood^10^. The sustained elevation in cardiovascular risk observed years after treatment suggests the presence of additional, possibly cumulative or delayed pathophysiological mechanisms that have yet to be fully elucidated.

While several studies have reported increased cardiovascular risk in CRC survivors^11,12^, relatively few have investigated the specific health of coronary arteries or performed direct coronary assessments. Most existing research relies on administrative data or surrogate markers of cardiovascular outcomes, with limited use of invasive techniques such as coronary angiography to assess the extent and nature of coronary artery disease. As a result, the direct impact of fluoropyrimidine-based chemotherapy on coronary artery integrity remains poorly characterized, and the burden of subclinical or undiagnosed coronary pathology in this population is likely underestimated.

This study aims to evaluate whether colorectal cancer survivors who are five years post-adjuvant chemotherapy with 5-fluorouracil (5-FU) or capecitabine exhibit evidence of coronary artery disease. By systematically characterizing cardiovascular outcomes in this cohort, we seek to elucidate the long-term cardiovascular impact of fluoropyrimidine-based chemotherapy and enhance our understanding of survivorship risks in the CRC population.

## Methods

### Study population

This cross-sectional, 2 year, single-center study was conducted at the University Clinical Center in Tuzla, at the Clinic for Invasive Cardiology and Clinic for Oncology and Radiotherapy (protocol summary in Figure 1). The study protocol was approved by the Ethics Committee of University Clinical Centre Tuzla (Ref. 02-09/2-40/24) and all methods were performed in accordance with their guidelines and regulations. Records of the 142 patients who received 5-fluorouracile (5-FU) and capecitabine adjuvant colorectal cancer chemotherapy between 1 January 2015 and 1 January 1 2018 were screened for inclusion in the study. Eligible patients (ChemT group n=45) were selected using the following inclusion criteria: (1) diagnosed with CRC as the primary and only tumour; (2) diagnosed between 2015 and 2018 (3) patients with CRC that received treatment with adjuvant ChemT (5-FU and capecitabine) for a minimum 5 years and maximum 7 years before the study started (Figure 1). The control group (n=45) was selected from 1245 patients scheduled for coronary angiography between 1 August 2021 and 1 May 2024. The eligible patients for the control group were selected using the following inclusion criteria: (1) cancer naïve, (2) age-matched to the CRC group (3) sex-matched to the CRC group (4) selected for elective coronary angiography.

**Figure 1.**
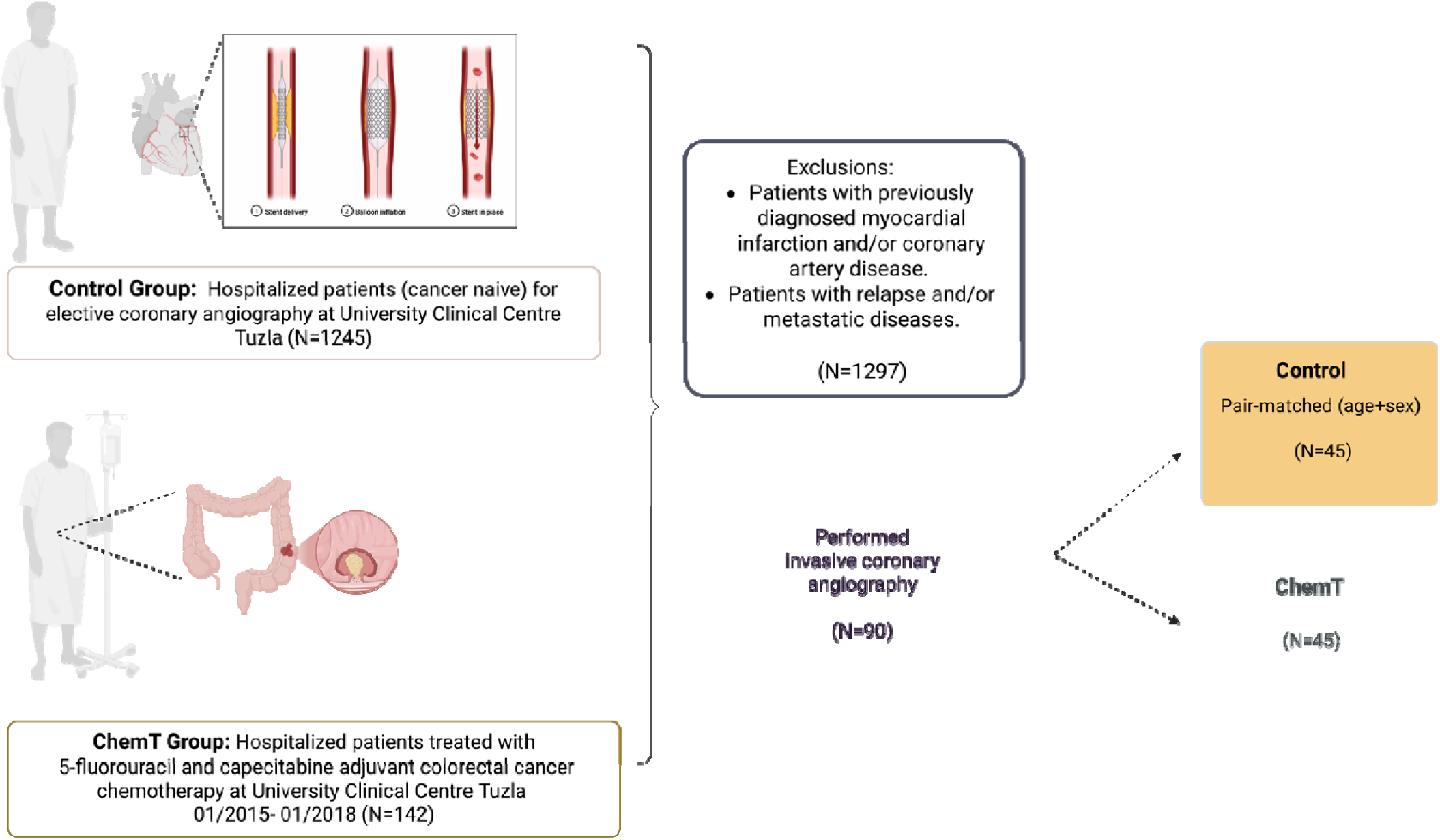
Summary of study design. Figure created with Biorender.com

Exclusion criteria were as follows: patients who did not give consent for the study, CRC patients with an initial diagnosis of metastatic cancer, those with cancer recurrence during the interval between completion of ChemT but prior to study enrolment, CRC patients who had additional ChemT beside 5-FU and capecitabine, patients with acute coronary syndromes or those previously diagnosed with CAD (Figure 1). We compared baseline clinical characteristics, treatment strategy and outcomes amongst two groups (ChemT vs Control). An overview of the patient cohort characteristics is shown in Table 1.

**Table 1.**
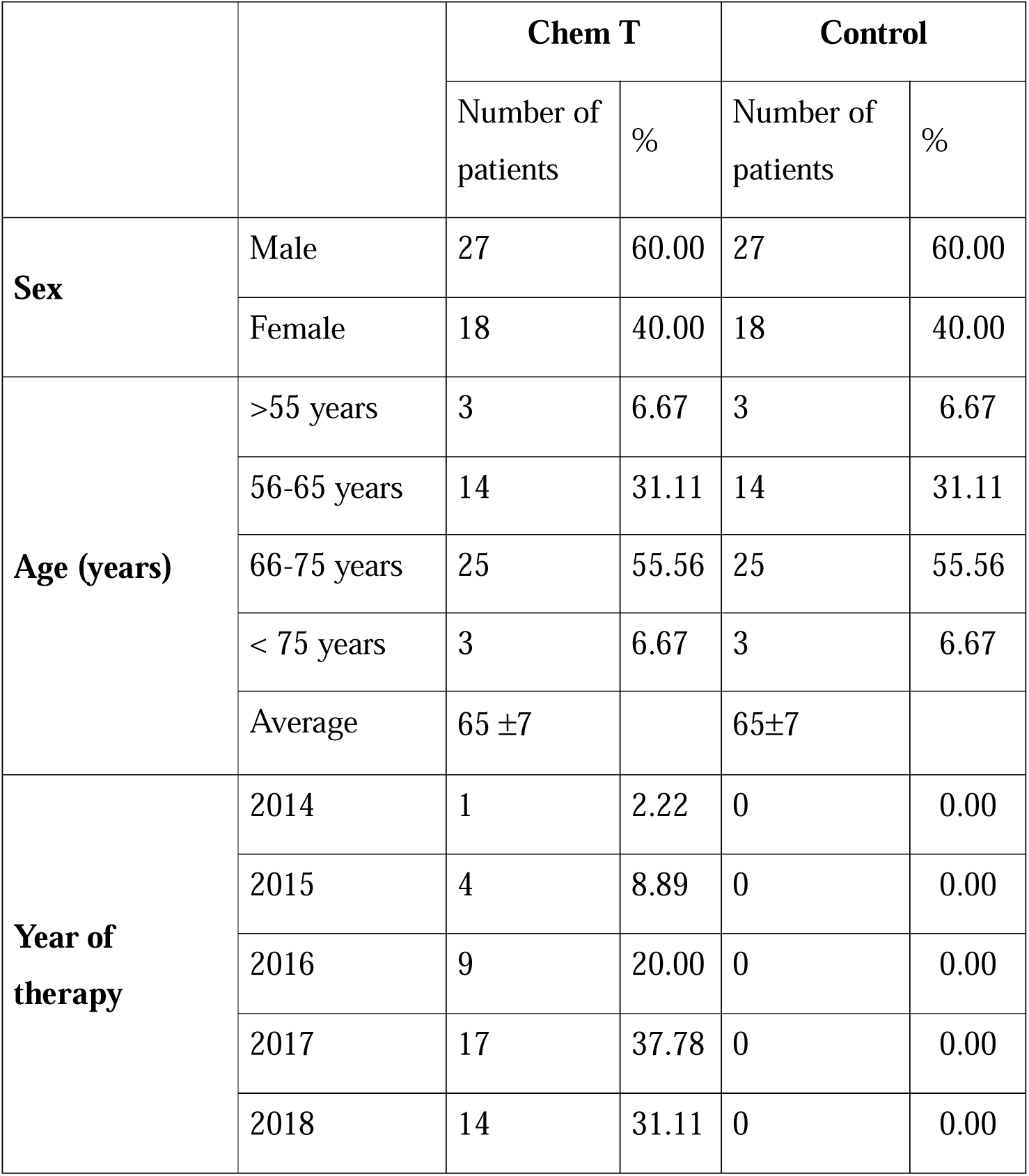
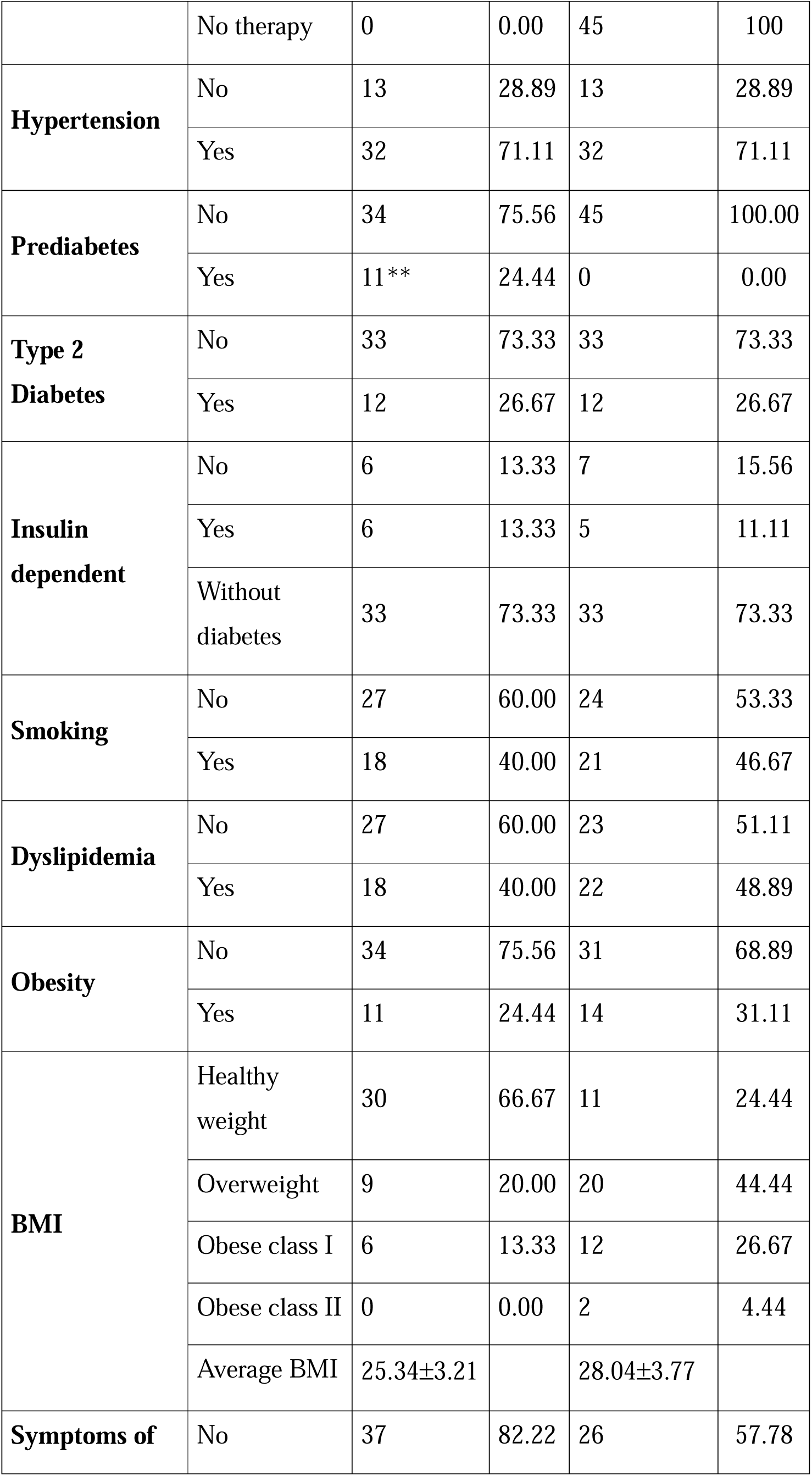
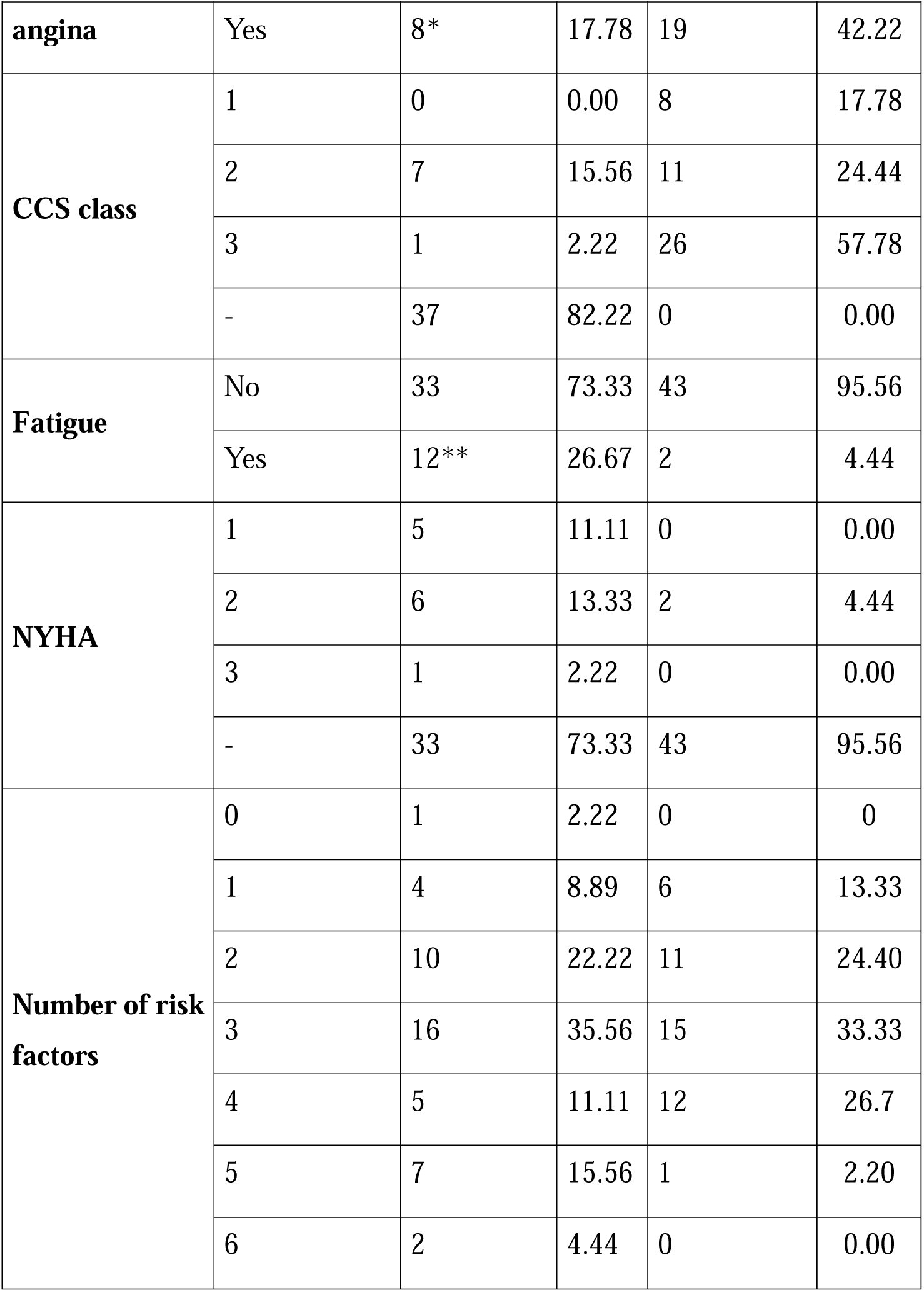
Patient characteristics at the time of hospitalization. Abbreviations: BMI-body mass index, CCS-Canadian Cardiovascular Society, NYHA-New York Heart Association. Data (qualitative variables) were compared with χ2 test. *P<0.05; **P<0.01

### Coronary Angiography and PCI Procedure

The coronary angiography procedure was performed with the adoption of a standard 5F-radial approach with a bolus of heparin (50 U/kg) and 200μg nitroglycerin in the radial artery. If the percutaneous coronary intervention (PCI) was needed and feasible during the index procedure, it was performed with the adoption of a standard radial approach that uses a 6F-guiding catheter. Procedures were performed by several experienced operators, each undertaking more than 250 PCI procedures per year. Coronary stenosis was estimated mostly visually, with a stenosis threshold of ≥50% for ostial and proximal lesions and ≥70% for mid and distal lesions.

After a bolus of heparin (100 U/Kg), 300 mg aspirin, and P2Y12 inhibitor in loading dose were given (clopidogrel 600 mg), target lesion was crossed through 0.014-inch wires. Non-ionic, low-osmolality contrast agent was used in all patients. After angioplasty, all patients were admitted to the Department of Interventional Cardiology. In addition, they were given 100 mg aspirin and 75 mg clopidogrel.

Concomitant medical treatment with β-blockers, angiotensin-converting enzyme inhibitors, and statins, were prescribed in accordance with the ESC Guideline for the management of the chronic coronary syndromes^13^. Echocardiography and antecubital venous blood samples for the laboratory analysis were collected before the index procedure.

### Statistical analysis

Data are presented as mean ± standard deviation (SD) unless otherwise stated. Normality of data distribution was examined using Shapiro–Wilk’s normality test. Comparison between two groups was performed by Student’s t-test (Gaussian data distribution). Qualitative variables were compared with Parson’s χ2 test and Fisher’s exact test. Statistical analysis was performed using SPSS software v.29 software. Differences were considered significant when P < 0.05.

## Results

The patient profile in terms of baseline demographic and cardiovascular risk characteristics for two cohorts: colorectal cancer (CRC) survivors treated with adjuvant fluoropyrimidine-based chemotherapy (Chem-T) and matched controls is summarized in Table 1. Both cohorts comprised 45 individuals with an identical sex distribution 60□% male and 40□% female. Age distribution was also comparable, with ∼87□% of participants in each group aged between 56 and 75 years. Chem-T group included patients treated from 2014 to 2018.

Comorbidities were frequent in both groups (Table 1). Hypertension was present in 71.11□% of patients in each cohort. Type 2 diabetes mellitus affected roughly one quarter of participants (26.67□%) in both groups, and the proportion requiring insulin was similar (13.33□% vs 11.11□%). However, prediabetes was recorded in 24.44□% of Chem-T patients and was absent among controls, suggesting a higher burden of glucose dysregulation in the treated cohort. Smoking prevalence was modestly higher in the control group (46.67□% vs 40.00□%), whereas dyslipidemia (48.89□% vs 40.00□%) and obesity (31.11□% vs 24.44□%) were also more common among controls. Body-mass index categories reflected these differences: the Chem-T cohort contained a greater proportion of participants with a healthy weight (66.67□% vs 24.44□%), whereas overweight and obesity (class I or II) were more prevalent in controls. Clinical manifestations differed between groups. Symptoms of angina were reported by 17.78□% of Chem-T survivors compared with 42.22□% of controls. Accordingly, Canadian Cardiovascular Society (CCS) class 3 angina (indicative of marked limitation of ordinary activity) was observed in only 2.22□% of Chem-T patients but in 57.78□% of controls. Fatigue was more prevalent in the Chem-T group (26.67□% vs 4.44□%), and this cohort also exhibited higher New York Heart Association (NYHA) functional class, with 11.11□% and 13.33□% of patients in classes I and II respectively. In contrast, 95.56□% of controls had no symptoms consistent with heart failure. The cumulative burden of cardiovascular risk factors was greater among Chem-T survivors. Only 2.22□% of treated patients had no traditional risk factors, compared with 0% of controls.

Conversely, 15.56□% of Chem-T patients had five risk factors and 4.44□% had six, whereas none of the controls had five or more. Together, these findings indicate that while many baseline characteristics were comparable between cohorts, CRC survivors exposed to fluoropyrimidine-based chemotherapy exhibited a higher burden of prediabetes, fatigue, NYHA functional limitation, and multiple cardiovascular risk factors, whereas untreated controls had higher rates of dyslipidemia, obesity, and angina symptoms. Participants in the CRC survivor group demonstrated significantly elevated aspartate aminotransferase (AST), alanine aminotransferase (ALT), and low-density lipoprotein (LDL) levels (Table 2). We have recently shown that CRC survivors have a long-term impairment in blood erythropoiesis^14^.

**Table 2.**
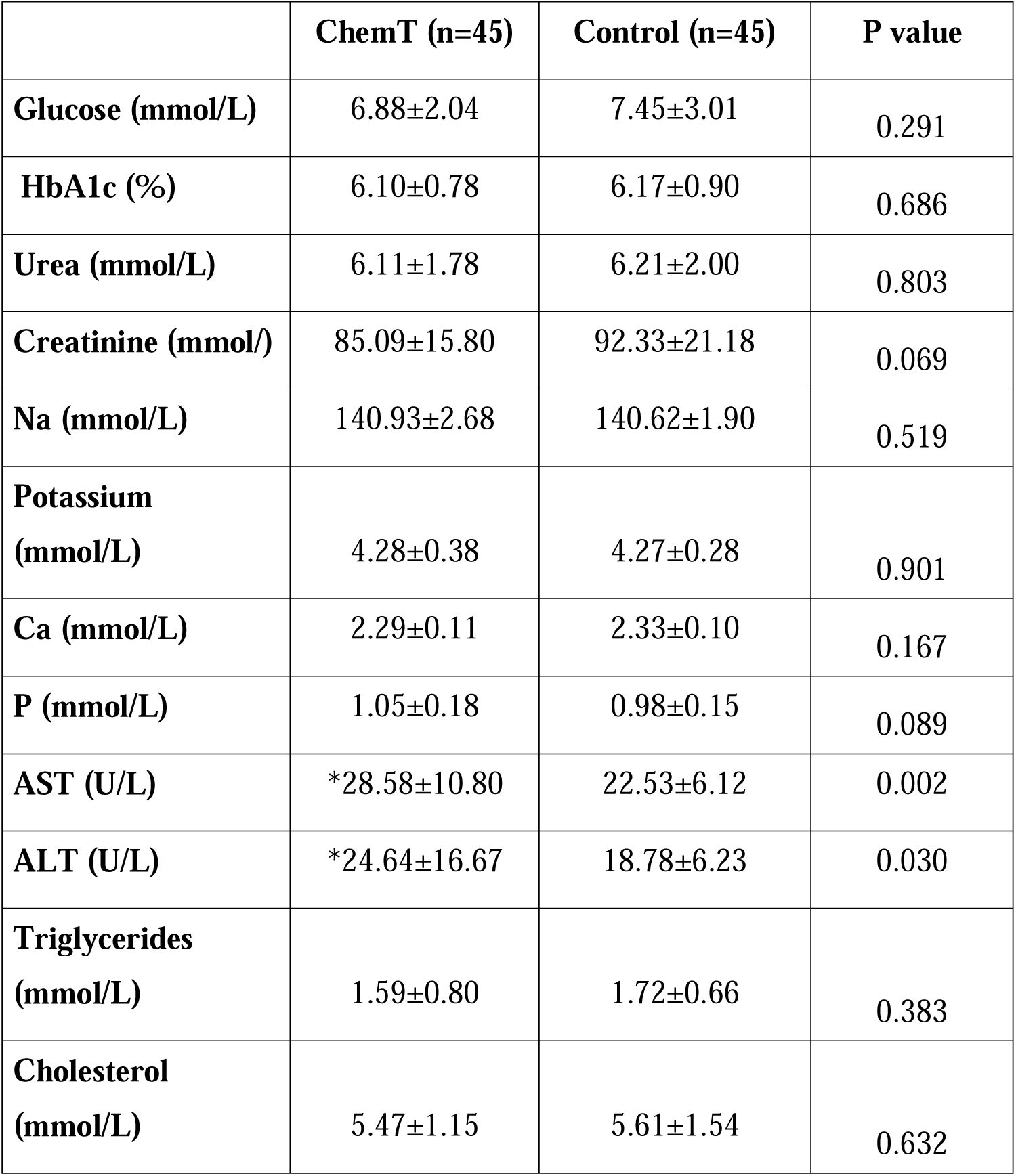
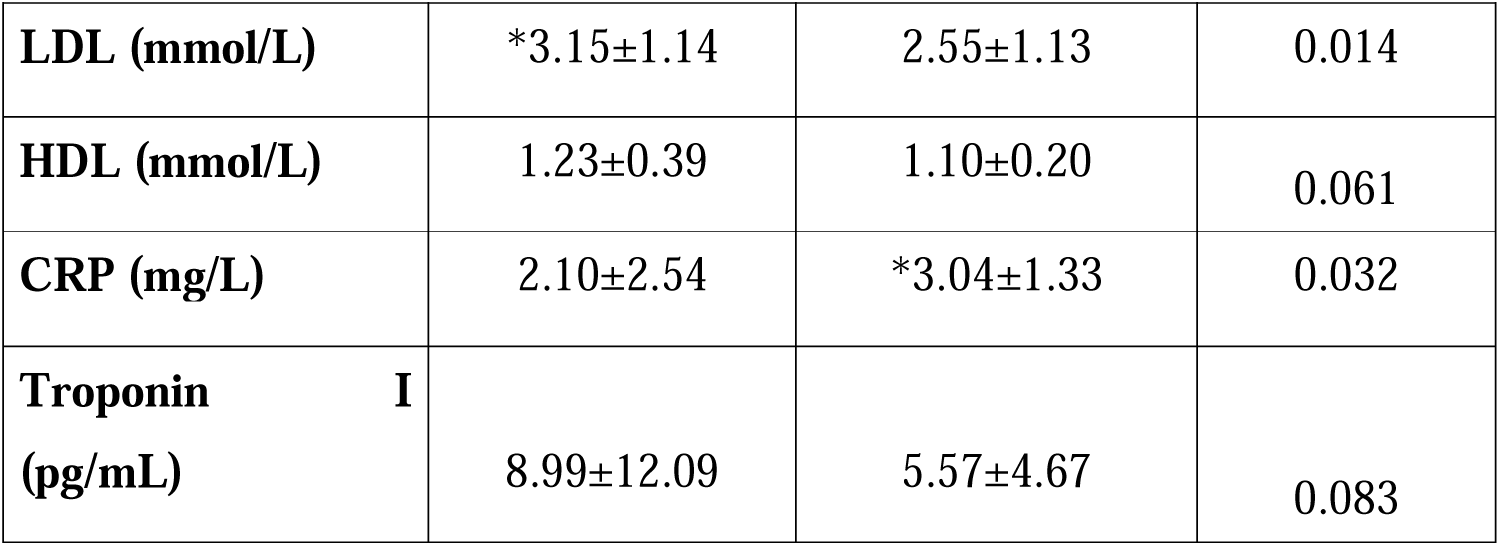
Patient plasma profile at the time of hospitalization. Data are presented as mean ± SD. Normality of data distribution was examined using Shapiro–Wilk’s normality test. Comparison between two groups was performed by Student’s t-test (Gaussian data distribution) or Mann-Whitney U test when data was non-normally distributed. *p<0.05 vs control. Abbreviations: HbA1c-Hemoglobin A1C, MCV – Mean Corpuscular volume; Na-Sodium, Ca – Calcium; P – Phosphorus; AST – Aspartate aminotransferase; ALT-Alanine aminotransferase; LDL – Low-density lipoprotein cholesterol; HDL-High-density lipoprotein cholesterol; CRP-C reactive protein;

|  | **ChemT (n=45)** | **Control (n=45)** | **P value** |
| --- | --- | --- | --- |
| **Glucose (mmol/L)** | 6.88±2.04 | 7.45±3.01 | 0.291 |
| **HbA1c (%)** | 6.10±0.78 | 6.17±0.90 | 0.686 |
| **Urea (mmol/L)** | 6.11±1.78 | 6.21±2.00 | 0.803 |
| **Creatinine (mmol/)** | 85.09±15.80 | 92.33±21.18 | 0.069 |
| **Na (mmol/L)** | 140.93±2.68 | 140.62±1.90 | 0.519 |
| **Potassium (mmol/L)** | 4.28±0.38 | 4.27±0.28 | 0.901 |
| **Ca (mmol/L)** | 2.29±0.11 | 2.33±0.10 | 0.167 |
| **P (mmol/L)** | 1.05±0.18 | 0.98±0.15 | 0.089 |
| **AST (U/L)** | *28.58±10.80 | 22.53±6.12 | 0.002 |
| **ALT (U/L)** | *24.64±16.67 | 18.78±6.23 | 0.030 |
| **Triglycerides (mmol/L)** | 1.59±0.80 | 1.72±0.66 | 0.383 |
| **Cholesterol (mmol/L)** | 5.47±1.15 | 5.61±1.54 | 0.632 |
| **LDL (mmol/L)** | *3.15±1.14 | 2.55±1.13 | 0.014 |
| **HDL (mmol/L)** | 1.23±0.39 | 1.10±0.20 | 0.061 |
| **CRP (mg/L)** | 2.10±2.54 | *3.04±1.33 | 0.032 |
| **Troponin I (pg/mL)** | 8.99±12.09 | 5.57±4.67 | 0.083 |

Echocardiographic evaluation (Table 3) revealed that patients in the control group had a statistically significant, although clinically minor, increase in left ventricular (LV) dimension; however, values remained within normal reference ranges in both groups. The control group also exhibited significantly greater interventricular septal thickness and a higher prevalence of concentric left ventricular hypertrophy.

**Table 3.** Transthoracic echo characteristics at the time of hospitalization. Data are presented as mean ± SD. Normality of data distribution was examined using Shapiro–Wilk’s normality test. Comparison between two groups was performed by Student’s t-test (Gaussian data distribution) or Mann-Whitney U test when data was nonnormally distributed. *p<0.05 vs control. Abbreviations:LVIDd -left ventricular internal dimension at end-diastole diameter: LAIDd - left atrium internal dimension diameter: IVSDd-Interventricular septum thickness at end/diastole; EFLV-left ventricular ejection fraction; LAVI-left atrial volume index; AV-aortic valve; PG-pressure gradient; PHT-pressure half time; MR-mitral regurgitation; TR-tricuspid regurgitation;

|  |  |  | P value |
| --- | --- | --- | --- |
|  | ChemT (n=45) | Control (n=45) |  |
| **LVIDd (cm)** | *4.81±0.49 | 5.12±0.47 | 0.003 |
| **LAIDd (cm)** | 3.84±0.41 | 3.80±0.95 | 0.795 |
| **IVSDd (cm)** | 1.16±0.15 | *1.42±0.79 | 0.037 |
| **EFLV (%)** | 55.73±6.50 | 55.96±6.71 | 0.874 |
| **LAVI (ml/m<sup>2</sup>)** | 30.20±8.00 | *33.28±5.83 | 0.040 |
| **Average E/e'** | 10.21±3.57 | 10.38±4.53 | 0.845 |
| **AV maximum velocity (m/sec)** | 1.58±0.66 | 1.71±0.61 | 0.315 |
| **AV mean PG (mmHg)** | 13.16±7.67 | 10.19±7.86 | 0.073 |
| **AV PHT (mmHg)** | 603.75±30.20 | 508.33±102.25 | 0.071 |
| **Vena contracta MR (cm)** | 1.59±1.50 | 1.82±2.07 | 0.547 |
| **EROA MR (cm<sup>2</sup>)** | 11.31±11.47 | 12.62±15.26 | 0.646 |
| **Vena contracta TR (cm)** | 2.58±1.31 | 2.99±1.83 | 0.407 |
| **EROA TR (cm<sup>2</sup>)** | 16.89±6.72 | 15.23±10.83 | 0.595 |
| **Ascending aorta (cm)** | 3.74±0.35 | 3.76±0.40 | 0.803 |

Coronary angiographic assessment of the left main coronary artery (LM) showed no statistically significant differences between the two groups regarding the presence of atherosclerotic disease overall or the presence of significant stenosis within any segment of the LM (Table 4).

**Table 4.** Coronary angiography results. Data are presented as numbers and percentage. Data (qualitative variables) were compared with χ2 test resulting in P values shown. Abbreviations: CAD-Coronary artery disease; LM-Left main; LAD-Left anterior descending artery: CX-Circumflex artery; RCA-Right coronary artery.

|  | **ChemT n (%)** | **Control n (%)** | **p value** |
| --- | --- | --- | --- |
| **Left Main (LM)** |  |  |  |
| Presence of any CAD in LM | 7 (15) | 2 (4) | 0.157 |
| Significant CAD in LM ostium | 0 (0) | 0 (0) | 0 |
| Significant CAD u LM shaft | 0 (0) | 0 (0) | 0 |
| Significant CAD in distal LM | 1 (2) | 1 (2) | 1.00 |
| **Left anterior descending artery (LAD)** |  |  |  |
| Presence of any CAD in LAD | 32 (71) | 20(44) | 0.019* |
| Significant CAD in LAD ostium | 3(6) | 1(2) | 0.616 |
| Significant CAD in proximal LAD | 11(24) | 1(2) | 0.004* |
| Significant CAD in mid LAD | 10(22) | 2(4) | 0.027* |
| Significant CAD in distal LAD | 0 (0) | 1 (2) | 1.000 |
| **Circumflex artery (CX)** |  |  |  |
| Presence of any CAD | 13 (28) | 11 (24) | 0.812 |
| Significant CAD in CX ostium | 0 (0) | 1 (2) | 1.000 |
| Significant CAD in proximal CX | 3 (7) | 1 (2) | 0.616 |
| Significant CAD in distal CX | 5 (11) | 1 (2) | 0.203 |
| **Right coronary artery (RCA)** |  |  |  |
| Presence of any CAD | 20 (44) | 15 (33) | 0.387 |
| Significant CAD in RCA ostium | 0 (0) | 0 (0) | 0 |
| Significant CAD in proximal RCA | 6(13) | 0(0) | 0.026* |
| Significant CAD in mid RCA | 7 (16) | 5 (11) | 0.756 |
| Significant CAD in distal RCA | 2 (4) | 1 (2) | 1.00 |

In contrast, the prevalence of atherosclerotic coronary artery disease in the left anterior descending artery (LAD) was significantly higher among participants in the CRC survivor group compared to controls (Figure 2). Moreover, a significantly greater number of participants had significant disease localized to the proximal and mid segments of the LAD (Figure 2). No significant differences were observed between groups in the prevalence of disease in the distal LAD or its diagonal branches (Table 4). There, were no statistically significant differences between groups in the presence or severity of atherosclerotic disease in the circumflex artery (CX) or its obtuse marginal (OM) branches (Table 4). However, a significantly greater proportion of patients in the CRC survivor group had significant disease in the proximal segment of the right coronary artery (RCA) compared to the control group (Figure 2). No significant differences were noted in the mid or distal segments of the RCA (Table 4).

**Figure 2.**
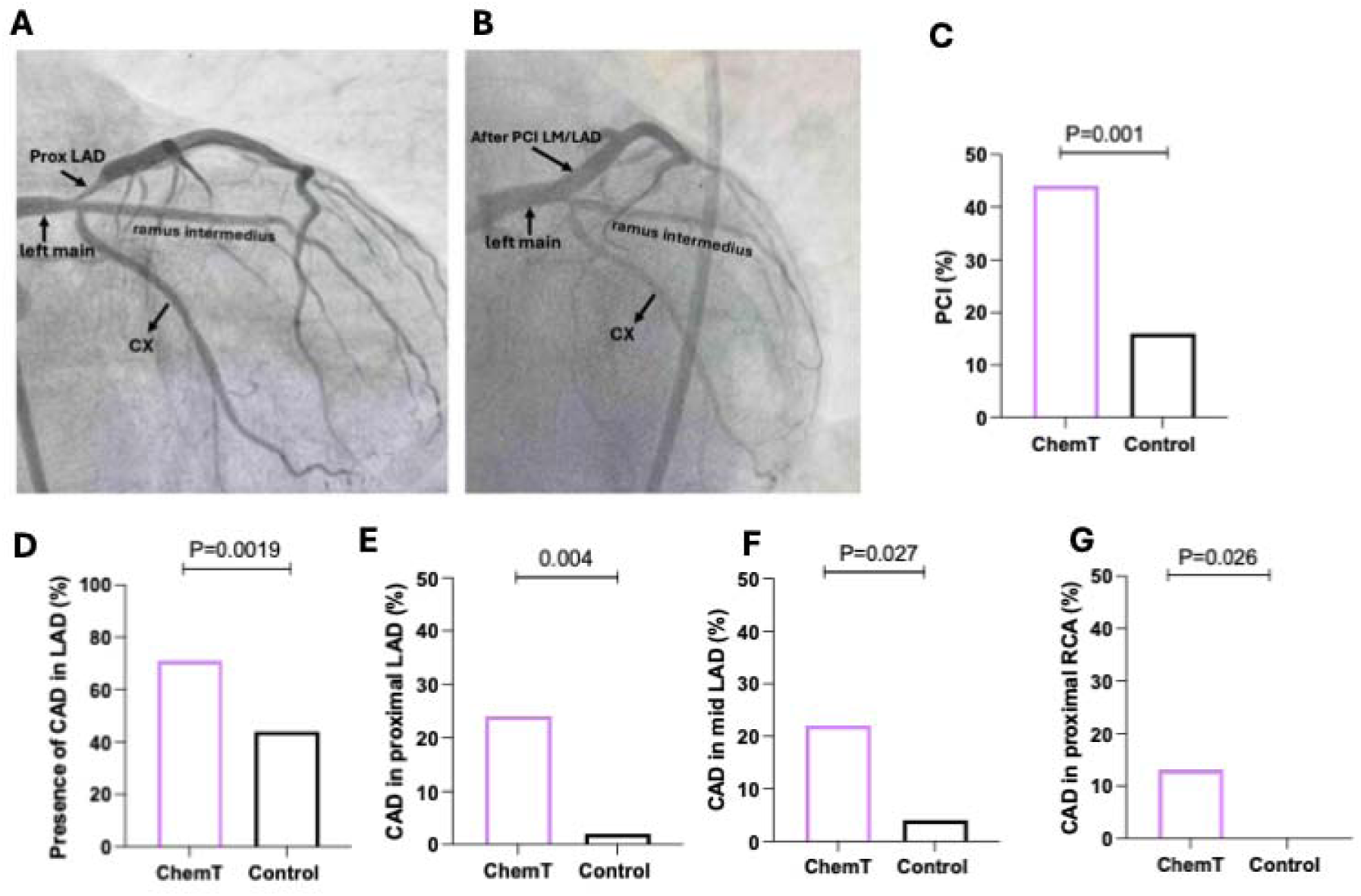
Prevalence of CAD was significantly higher in ChemT patients. (**A**), (**B**)Representative angiograms of female study participant in her 50s, hypertension, without typical anginal symptoms, received 5-FU seven years prior to the index procedure. (**A**) Before PCI (**B**) after PCI. LM/prox LAD with DES 4.0 x 26mm and POT with NC 4.5 x12mm. (**C**) Percentage of PCI procedures per group (**D**) presence of any CAD in LAD (**E**) significant CAD in prox LAD (**F**) significant CAD in mid LAD (**G**) significant CAD in prox RCA. Data (qualitative variables) were compared with χ2 test resulting in P values shown. Data in plots are presented as percentage of total number of patients (n=45/group).

Regarding revascularization strategies (Tables 5,6), a significantly higher number of participants in the CRC survivor group underwent percutaneous coronary intervention (PCI, Figure 2), whereas significantly more patients in the control group were managed with medical therapy alone (Table 5). One patient in each group underwent coronary artery bypass grafting (CABG). Furthermore, the total number of coronary stents placed was significantly greater in the CRC group. However, there were no significant differences between groups in either the maximum stent diameter or the cumulative stent length (Tables 5,6).

**Table 5.**
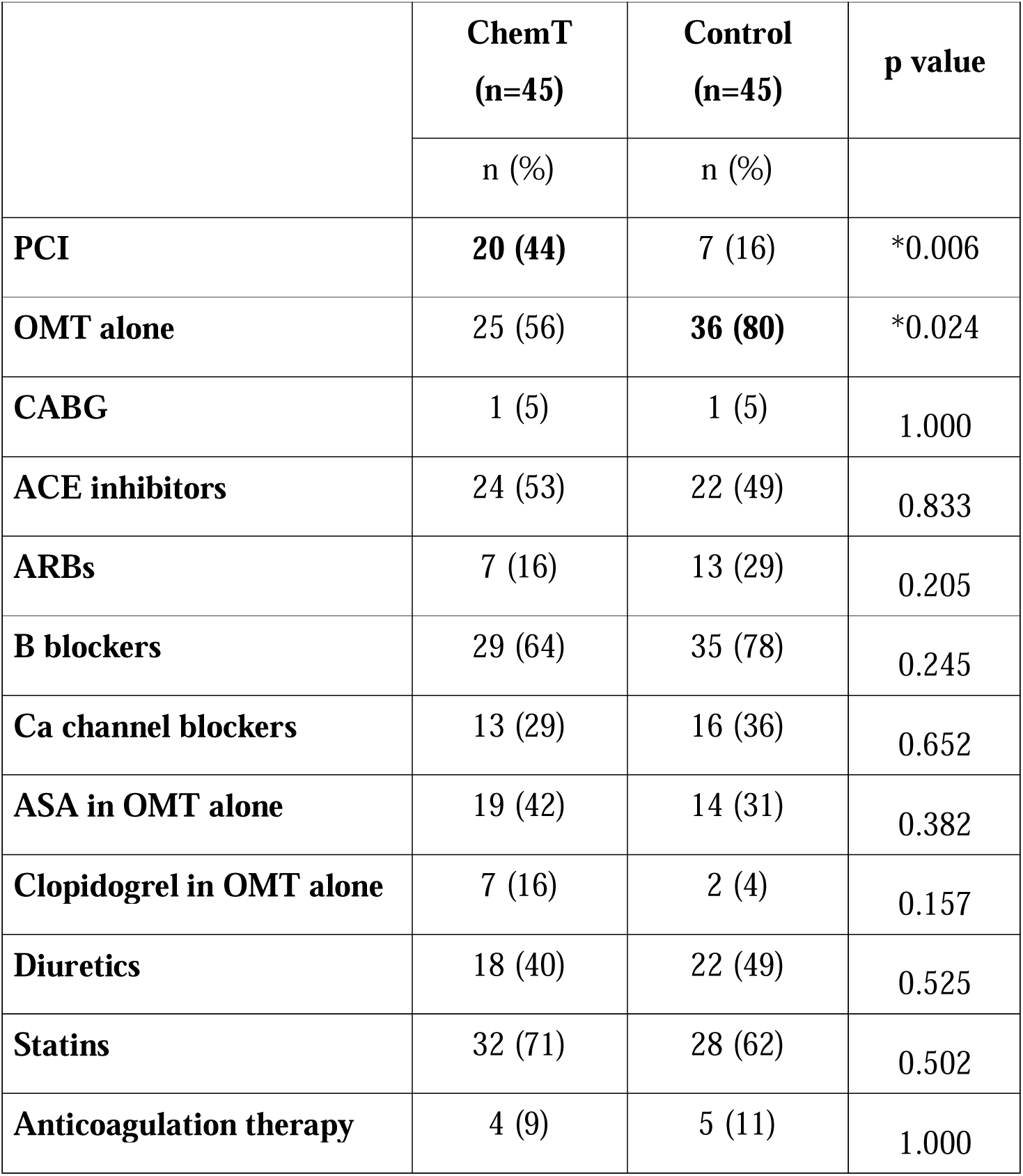
Treatment details. Data are presented as numbers and percentage. Data (qualitative variables) were compared with χ2 test resulting in P values shown. Abbreviations: PCI-percutaneous coronary intervention; OMT-optimal medical treatment; CABG-coronary artery bypass graft; ACE-angiotensin converting enzyme; ARBs-angiotensin receptor blockers; B-beta; Ca-Calcium; ASA-acetylsalicylic acid.

**Table 6.** Treatment details. Data are presented as numbers, percentage. Data (qualitative variables) were compared with χ2 test resulting in P values shown. Lesion charteristics data presented as mean ± SD. Normality of data distribution was examined using Shapiro–Wilk’s normality test and comparison between two groups was performed by Student’s t-test (Gaussian data distribution). Abbreviations: PCI-percutaneous coronary intervention; SC-semi-compliant; DCB-drug coating balloon; NC-non-compliant.

|  | **ChemT** |  | **Control** |  | **p value** |
| --- | --- | --- | --- | --- | --- |
|  | Population size (n) | n (%) | Population size (n) | n (%) |  |
| **Calcified lesions** | 20 | 10 (50) | 7 | 1 (14) | 0.183 |
| **DCB** | 20 | 4 (20) | 7 | 0 (0) | 0.545 |
| **NC balloon post-dilatation** | 20 | 11 (55) | 7 | 3 (43) | 0.678 |
| **Procedure complications** | 20 | 1 (5) | 7 | 0 (0) | 1.000 |
| **Death** | 20 | 0 (0) | 7 | 0 (0) | 0 |
| **Lesion characteristics** |  |  |  |  |  |
| **Total stent length (mm)** | 18 | 29.44±11.54 | 7 | 26.86±9.65 | 0.605 |
| **Maximal stent diameter (mm)** | 18 | 3.64±0.50 | 7 | 3.57±0.34 | 0.728 |
| **Total DCB length (mm)** | 4 | 23.00±3.83 | 0 | - | - |
| **Maximal DCB diameter (mm)** | 4 | 2.81±0.239 | 0 | - | - |

## Discussion

This cross-sectional, angiography-based study provides novel evidence that colorectal cancer (CRC) survivors treated with adjuvant fluoropyrimidine-based chemotherapy, specifically 5-fluorouracil (5-FU) or capecitabine, have a significantly greater burden of coronary artery disease (CAD) compared with age- and sex-matched cancer-free controls. The coronary disease was most prominent in the proximal segments of the left anterior descending (LAD) and right coronary artery (RCA) anatomical regions known for their prognostic importance in cardiovascular outcomes. These findings suggest that long after completion of chemotherapy, survivors remain at elevated risk for silent, yet clinically significant, CAD, reinforcing a critical gap in survivorship care.

### Long-Term Cardiovascular Sequelae of Fluoropyrimidines

Acute cardiotoxicity from 5-FU and capecitabine ranging from chest pain to myocardial infarction is well documented, with incidence rates of 1%–4%^8,15^. However, the long-term cardiovascular consequences have remained insufficiently characterised. Prior registry-based studies report cardiovascular mortality as a major cause of death among CRC survivors, with a 10-year CVD incidence approaching nearly threefold that of the general population^7,16^. Yet, few investigations have directly evaluated coronary anatomy or assessed angiographic outcomes in this population.

Our study is among the first to provide invasive imaging evidence of fluoropyrimidine-associated coronary pathology. We identified a significantly higher prevalence of anatomically significant stenoses in proximal coronary segments 71% of chemotherapy-treated patients had proximal LAD disease, compared with 44% of controls; and 13% had proximal RCA disease, compared to none in the control group. Given that the proximal LAD supplies a large myocardial territory and is strongly associated with major adverse cardiovascular events^17,18^, these findings suggest that fluoropyrimidine therapy may predispose survivors to high-risk, clinically silent coronary phenotypes.

### Mechanistic Insights: Structural and Haematologic Contributions

The pathophysiological mechanisms driving these findings are likely multifactorial. Fluoropyrimidines have been shown to induce direct endothelial injury via oxidative stress, nitric oxide depletion, and inflammation, as well as provoke coronary vasospasm and promote vascular smooth muscle proliferation^19,20^. Capecitabine and 5-FU have been associated with both functional (vasospastic) and structural (atherosclerotic) coronary abnormalities, which may persist long after therapy cessation^21^. These processes create a vascular environment conducive to accelerated atherogenesis, particularly in vulnerable proximal segments^22^.

In parallel, our cohort demonstrated systemic haematologic and biochemical alterations consistent with a pro-atherogenic milieu. Chemotherapy-exposed patients had significantly lower serum iron and erythrocyte counts, along with elevated mean corpuscular haemoglobin concentration (MCHC)^14^. These changes are associated with impaired oxygen delivery, increased blood viscosity, and microvascular dysfunction all of which may contribute to coronary ischaemia. Iron, a critical cofactor in mitochondrial energetics and vascular homeostasis, has been independently linked to adverse cardiovascular outcomes^22^. Thus, its deficiency may represent both a marker and mediator of vascular risk in this population.

Elevated LDL-cholesterol levels observed in chemotherapy-treated patients further compound cardiovascular risk. Although the relationship between LDL and CRC remains inconclusive, LDL is a key driver of atherogenesis, and chemotherapy may shift lipid profiles toward more atherogenic subtypes, including oxidized LDL, which exacerbates endothelial dysfunction and plaque instability^23,24^.

### Clinical Expression and Interventional Implications

Despite having fewer self-reported symptoms of angina, chemotherapy-exposed patients were more likely to undergo percutaneous coronary intervention (PCI) compared to controls (44% vs 16%). They also received more coronary stents, underscoring the clinical severity and procedural implications of their disease. This discrepancy between symptomatic presentation and objective disease burden highlights a potentially silent progression of CAD possibly due to altered pain perception, autonomic dysfunction, or reduced physical activity in cancer survivors^25,26^.

The anatomical pattern of disease concentrated in high-risk proximal segments emphasizes the need for proactive cardiovascular surveillance. Functional testing alone may fail to detect subclinical but clinically relevant disease in this population, reinforcing the potential value of anatomical imaging in selected high-risk survivors.

Our findings suggest several important directions for clinical practice and future investigation. Routine monitoring of simple laboratory biomarkers including serum iron levels, complete blood count indices, and lipid profiles may offer a practical and early approach to identifying CRC survivors at elevated cardiovascular risk. These parameters, which reflect iron status, hematologic health, and lipid metabolism, could serve as accessible tools for risk stratification during survivorship follow-up. In parallel, the high burden of coronary artery disease observed in this population underscores the need for preventive strategies aimed at aggressive risk factor modification. This may include the early initiation of lipid-lowering therapies, antiplatelet regimens, and potentially iron repletion, each of which warrants prospective evaluation in dedicated clinical studies. Additionally, mechanistic research is essential to elucidate the vascular consequences of fluoropyrimidine chemotherapy. Future studies should investigate the molecular pathways of endothelial dysfunction, chronic inflammation, and chemotherapy-induced lipid dysregulation. Longitudinal imaging studies and serial biomarker analyses will be critical to better understand the progression of coronary disease in cancer survivors and to guide targeted interventions.

This study has several limitations. It was conducted at a single center with a moderately sized cohort, which may limit generalizability. Although groups were closely matched by age and sex, residual confounding cannot be fully excluded, particularly given baseline differences in cardiovascular risk profiles. The cross-sectional design also precludes causal inference. In addition, plaque morphology and coronary microvascular dysfunction were not assessed. The presence and severity of coronary lesions were assessed by visual estimation by experienced operators rather than advanced imaging techniques, as there were no true bifurcation lesions (Medina 1,1,1) or left main bifurcations. While our findings demonstrate an association between ChemT exposure and increased atherosclerosis burden, we cannot determine whether this relationship is mediated through ChemT-induced changes in traditional cardiovascular risk factors or reflects underlying baseline differences. Prospective studies with detailed cardiovascular risk factor assessment, longitudinal follow-up, and comprehensive adjustment using multivariable modeling are needed to clarify these pathways.

### Conclusions

CRC survivors exposed to fluoropyrimidine-based chemotherapy have a significantly higher burden of obstructive coronary artery disease, particularly in anatomically high-risk segments of the LAD and RCA. This finding, coupled with systemic biochemical and hematologic alterations, supports a multifactorial model of delayed vascular toxicity. As cancer survivorship continues to improve, our results underscore the urgent need to expand the cardiovascular lens of oncology follow-up transforming survivorship care into a truly multidisciplinary endeavour.

## Data Availability

All data produced in the present work are contained in the manuscript

## Acknowledgements

The authors express their gratitude to all patients who participated in the study. We acknowledge collegiate support from Ms Sanela Idrizovic, University Clinical Centre Tuzla. DA acknowledges funding from the Wellcome Trust (221604/Z/20/Z) and Barts Charity (G-002145).

